# Prevalence and Management of Chronic Kidney Disease in Irish Primary Care Network

**DOI:** 10.64898/2026.10.27.26366639

**Authors:** Benjamin M. Jacob, Keelan Eyers, Danyao Jin, Michael Hayes, Aoife Coughlan, Emma Ryan, Benjamin van Hecke, Patrick Redmond, Austin G. Stack, Donal Sexton, Donal Bailey

**Author notes:** **Corresponding author:** Benjamin M. Jacob.

## Abstract

**Background:** Chronic kidney disease (CKD) imposes a substantial burden on health systems and society. Although early detection and guideline-recommended management can improve outcomes, opportunities remain to enhance management in primary care.

**Aim:** To evaluate CKD prevalence, guideline-directed medical therapy (GDMT), and monitoring in Irish primary care.

**Design and Setting:** Retrospective cohort study using community-based electronic health records (EHR) from the CRADLE database, across 66 Irish practices.

**Methods:** Active adults aged ≥20 years on 30 July 2025 were included. CKD prevalence was determined using estimated glomerular filtration rate (eGFR) and/or urine albumin-to-creatinine ratio (uACR). Descriptive analyses of epidemiology, prescribing, and monitoring were conducted.

**Results:** Of 468,534 adults, 19,382 met the criteria of laboratory-confirmed CKD: age- and sex-standardised prevalence was 5.36%. 74.0% of laboratory-confirmed CKD patients have ≥1 cardiometabolic comorbidities. Within 12 months of laboratory-confirmed CKD, monitoring increased over time, with 66.0% of patients receiving follow-up eGFR test and 15.1% receiving uACR test, falling to 6.6% among patients without type 2 diabetes (T2DM). Among GDMT-eligible CKD patients, 64.0% received any renin-angiotensin-aldosterone inhibitors (RAASi) and 30.9% received any sodium-glucose cotransporter 2 inhibitors (SGLT2i) after they met their prescription eligibility for CKD, but SGLT2i use dropped to 9.4% among those without T2DM or heart failure.

**Conclusion:** CKD and cardiometabolic comorbidities impose a substantial burden in Irish primary care. Despite improved kidney function testing, albuminuria testing remained low, particularly in non-diabetic patients, and GDMT use was suboptimal. Improving albuminuria testing and timely GDMT offers important opportunities to improve CKD outcomes in Irish primary care.

## Introduction

Chronic kidney disease (CKD) affects approximately 10–13% of adults worldwide.^1,2^ Early detection and treatment can slow progression, reduce cardiovascular risk and premature mortality, and lower costs.^1,3–8^ Reliable population-level data are essential for planning and evaluating the quality of CKD management in primary care.^1,3^

In Ireland, prevalence estimates range from 4–5% in the general population to approximately 16% in adults ≥50 years, with most cases occurring in later life.^9–12^ Evidence from The Irish Longitudinal Study on Ageing (TILDA) suggests most of individuals with CKD are unaware, reflecting its largely asymptomatic nature in early stages.^9–11^

Existing evidence comes from research cohorts and routine health data. Cohort studies benefit from systematic testing but may not fully represent the broader population. Conversely, routine data can underestimate undiagnosed CKD while providing insight into the real-world recognised burden.^9–12^ These findings highlight an opportunity to improve CKD identification, earlier diagnosis and timely management in Irish primary care.^13^

Effective CKD management relies on early detection, regular monitoring, and use of GDMTs, including RAASi and SGLT2i which can slow disease progression.^14^ Regular monitoring of kidney function and albuminuria is essential for risk assessment and treatment decisions,^15^ yet albuminuria testing is not consistently performed in primary care, limiting early detection and accurate risk stratification.^16^

Despite established guidelines, important gaps persist in CKD diagnosis, monitoring and GDMT use, including opportunities to improve diagnosis and management.^16–19^ Large-scale evaluations in Irish primary care are limited. Evidence suggests that earlier identification and timely intervention, including appropriate treatment and follow-up, can improve patient outcomes and support more efficient use of resources.^20^

Using Irish primary care EHR, this study provides the first large-scale assessment of CKD burden and monitoring practices, identifying opportunities to improve albuminuria monitoring and optimise GDMT uptake.

## Methods

### Study design and setting

This retrospective cohort study used routinely collected primary care EHR data, from the Clinical Research and Data-Driven Learning Environment (CRADLE), which contains data from Centric Health general practices across Ireland. Records from January 2006, or each patient’s earliest entry, to the index date (30 July 2025) were analysed to assess

CKD prevalence, epidemiology, management, and monitoring. Data included diagnoses, consultations, prescriptions, laboratory results, and clinical correspondence.

### Participants

Patients were classified as having CKD if they had either a CKD diagnostic code or laboratory evidence of CKD. Three definitions were applied (Supplementary Table 1). The strict definition approximated KDIGO chronicity criteria, requiring two eGFR measurements <60 mL/min/1.73 m^2^ at least 90 days apart within 12 months, and/or persistent albuminuria defined by one uACR >70 mg/mmol or two uACR values of 3-70 mg/mmol at least 90 days apart. The practical definition, used for primary analyses, retained the eGFR criterion but broadened albuminuria identification to include a single uACR >3 mg/mmol. The expanded definition, used for sensitivity analyses and comparison with TILDA, additionally included patients with a single most recent eGFR <60 mL/min/1.73 m^2^ without a preceding normal result in the prior 12 months, as well as those with a coded CKD diagnosis.

### Variables and outcomes

Variables included demographic, laboratory, comorbidity, prescribing, and monitoring data. The CKD Practical definition was used for primary analyses and the Expanded definition for sensitivity analyses and comparison with TILDA. Cardiometabolic comorbidities (hypertension, T2DM, cardiovascular disease, and heart failure) were identified from diagnostic codes (Supplementary Table 2). Prescribing data assessed eligibility and initiation of RAASi and SGLT2i therapies (Supplementary Table 3). Laboratory data were used to evaluate the timing of eGFR and uACR monitoring after CKD identification.

### Data Cleaning

Laboratory data were harmonised across practices by standardising eGFR and uACR test labels and converting uACR values to mg/mmol where required.

### Statistical analysis

Analyses were conducted within the secure CRADLE environment using Python and PySpark, with 95% confidence intervals calculated using binomial methods. CKD prevalence was estimated using three definitions, with the practical definition used for primary analyses, and results were stratified by age, sex, and cardiometabolic comorbidity, with standardisation to the Irish adult population.^21^

Prescribing and monitoring were assessed at the index date and following CKD identification. Analyses examined treatment initiation following eligibility and the timing of initiation, as well as follow□up testing of eGFR and uACR, including within 12 months of laboratory-confirmed CKD. Timely prescribing (≤6 months) and timely monitoring (≤12 months) were examined using Poisson regression with robust variance estimators, adjusting for age, sex, and cardiometabolic comorbidities.

### Sensitivity Analyses

Prevalence was re-estimated using a denominator restricted to tested patients and using the Strict and Expanded CKD definitions. Prescribing and monitoring models were repeated in subgroups with ≥6 months and ≥12 months of prior follow-up, respectively.

## Results

### CKD Prevalence

By 30 July 2025, 19,382 of 468,534 active adults (≥20 years) in the CRADLE database, met the practical laboratory definition of CKD and formed the laboratory-confirmed cohort (Supplementary Figure 1).

Under the practical definition, the crude prevalence of CKD was 4.1% in adults ≥20 years, rising to 5.4% after age- and sex-standardisation. Across the three laboratory-based definitions, standardised prevalence ranged from 4.8% to 6.2%. When analyses were restricted to individuals who underwent kidney testing, standardised prevalence increased to 7.1%. Among adults ≥20 years, crude prevalence was 8.8% in the tested population compared with 4.1% when using the full denominator; among those aged ≥50 years, corresponding crude estimates were 15.0% versus 10.1%. After standardisation, prevalence remained higher in tested individuals (7.1% vs 5.4% in adults ≥20 years; 14.6% vs 11.4% in those ≥50 years) (Table 1).

**Table 1.** Crude and age–sex standardised prevalence of CKD across three operational definitions.

|  | Main Analyses | Sensitivity Analyses |  |  |
| --- | --- | --- | --- | --- |
|  | Practical CKD | Strict CKD | Expanded CKD | Practical CKD in individuals with kidney function test |
| | Overall study population aged $\geq 20$ years (N = 468,534) | | | |
| Crude | 4.1% | 3.6% | 4.8% | 8.8% |
| Standardised | 5.4% | 4.8% | 6.2% | 7.1% |
| | Subgroup aged $\geq 50$ years (N = 181,302) | | | |
| Crude | 10.1% | 9.1% | 11.6% | 15.0% |
| Standardised | 11.4% | 10.3% | 13.0% | 14.6% |

CKD: chronic kidney disease.
Denominator restricted to tested patients only. To assess the impact of limiting the denominator to tested patients, we compared CKD prevalence estimates based on patients who had undergone an eGFR or uACR test with estimates derived from the full active practice population.

CKD prevalence increased markedly with age, from 0.1% in adults aged 20–29 years to 38.7% in those ≥80 years (Supplementary Figure 2; Supplementary Table 4). Among individuals ≥ 50 years, the standardised prevalence was 11.4% under the practical definition and 14.6% when restricting to individuals with kidney testing (Table 1).

### Baselines characteristics and comorbidities

Table 2 summarises the characteristics of the active CRADLE population aged >20 years (N=468,534) and the practical CKD subpopulation (N=19,382). Compared with the overall population, patients meeting the practical CKD criteria were substantially older (mean age of 73.3 and median age of 76 years ([IQR 66-82]), versus 46.3 and 44 years [IQR 33-57], respectively). The proportion of males was similar (44.3% in the CKD subpopulation vs 45.8% in the active population).

**Table 2.**
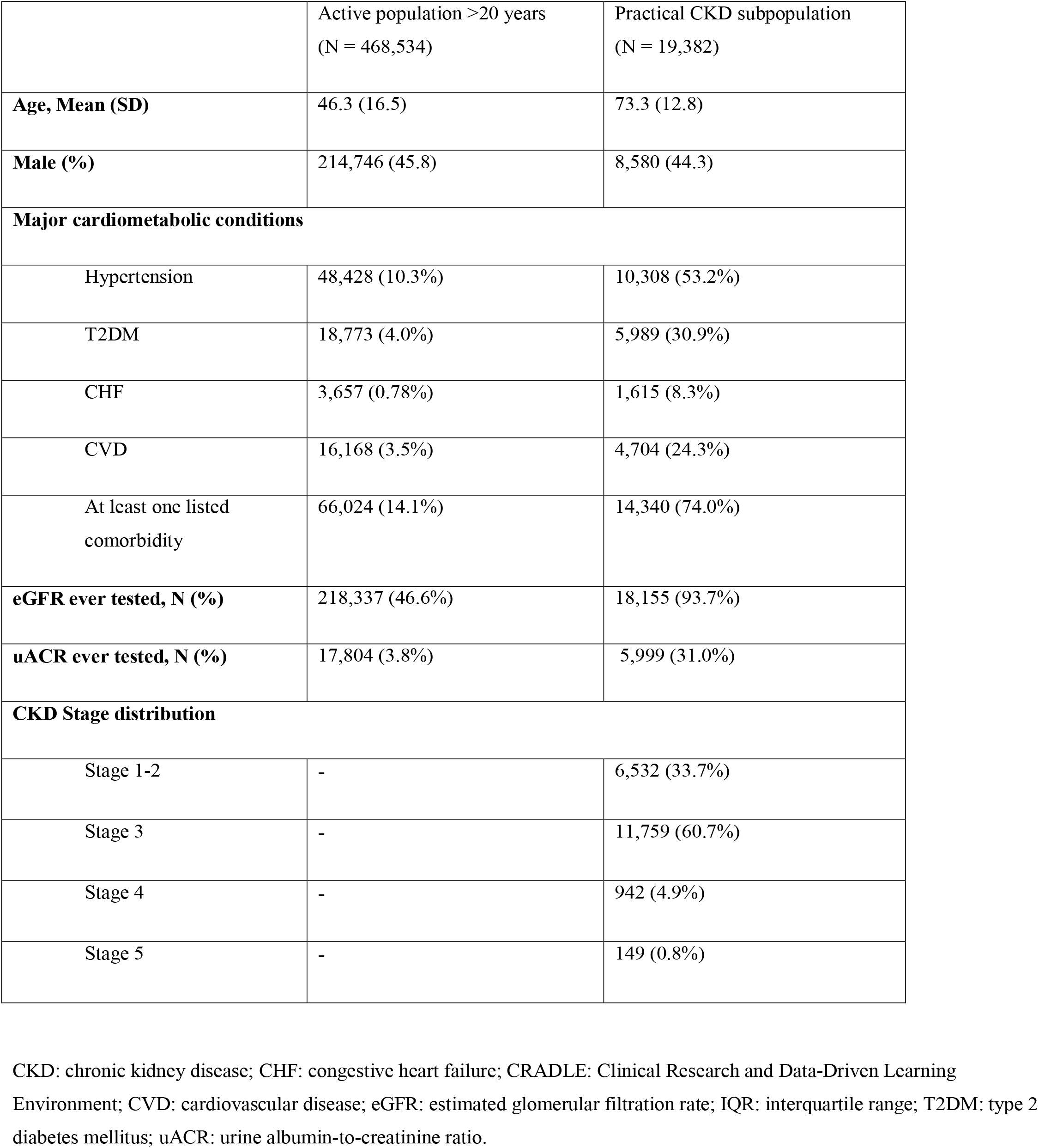
Demographic characteristics, laboratory values, stage distribution and comorbidities of the patients of the active population of the CRADLE database and the patients who met CKD criteria (Practical CKD subpopulation).

CKD patients had a markedly higher cardiometabolic burden. Hypertension was present at 53.2%, T2DM at 30.9%, cardiovascular disease (CVD) at 24.3%, and chronic heart failure (CHF) at 8.3%, compared with 10.3%, 4.0%, 3.5%, and 0.8%, respectively, in the overall population Overall, 74.0% of CKD patients had at least one listed comorbidity, compared with 14.1% in the overall population (Table 2).

Among CKD patients, 93.7% had undergone eGFR testing, and 31.0% had received a uACR test. In the overall population, testing rates for eGFR and uACR were far lower (46.6% and 3.8%, respectively). Most patients with laboratory-confirmed CKD were classified as having CKD stage-3 (60.7%), followed by 33.7% in stage 1-2 (Table 2).

### Cohort follow-up and monitoring practices

EHR coverage was extensive: median follow-up was 2.7 years (IQR 1.3-6.1 years) before the practical CKD definition, and 2.2 years afterwards.

Within 12 months of laboratory-confirmed CKD, 66.0% of patients received follow-up eGFR testing, whereas only 15.1% underwent uACR testing. Patients with T2DM had higher testing rates for HbA1c (77.0%) and eGFR (71.9%), and uACR (34.2%) (Figure 1A). In contrast, for patients without T2DM, eGFR and HbA1c testing remained common (63.4% and 57.9%, respectively), but uACR testing was infrequent (6.6%) (Figure 1B).

**Figure 1.**
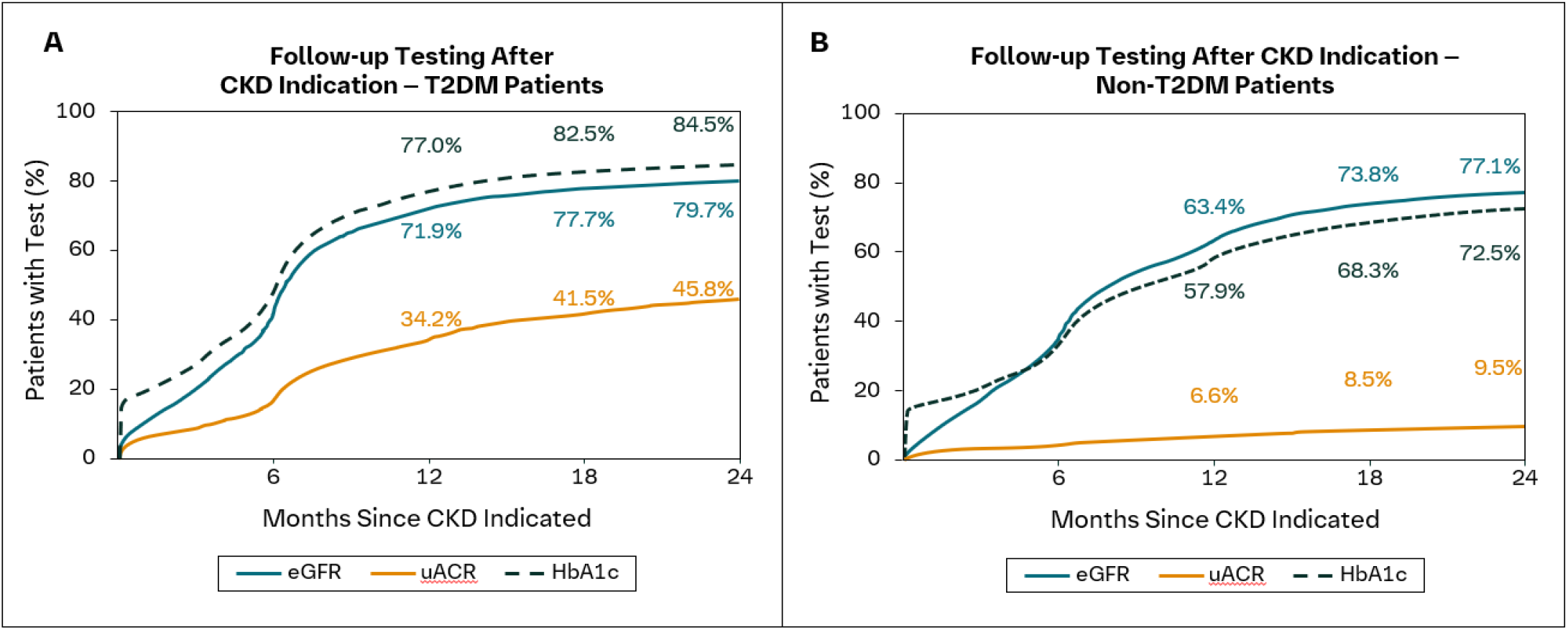
Time to First Follow-Up Testing Post CKD Indication for eGFR, uACR, and HbA1c in Patients With T2DM (A) and Without T2DM (B). CKD: chronic kidney disease; eGFR: estimated glomerular filtration rate; HbA1c: glycated haemoglobin; T2DM: type 2 diabetes mellitus; uACR: urine albumin-to-creatinine ratio.

### Prescriptions of GDMT

Among eligible patients, 64.0% received at least one RAASi prescription on or after their eligibility date, while 30.9% received a SGLT2i after meeting eligibility criteria. No nsMRA prescriptions were identified. Prescribing rates for RAASi and SGLT2i were generally higher among patients with comorbidities providing additional treatment indications. For RAASi, prescribing ranged from 67.9% among patients with T2DM to 69.4% among those with CVD, compared with 46.7% among patients without CHF or hypertension. For SGLT2i, prescribing rates were highest among patients with CHF (46.6%), followed by T2DM (41.1%) and CVD (35.7%), and lowest among those without T2DM or CHF (9.4%) (Table 3).

**Table 3.**
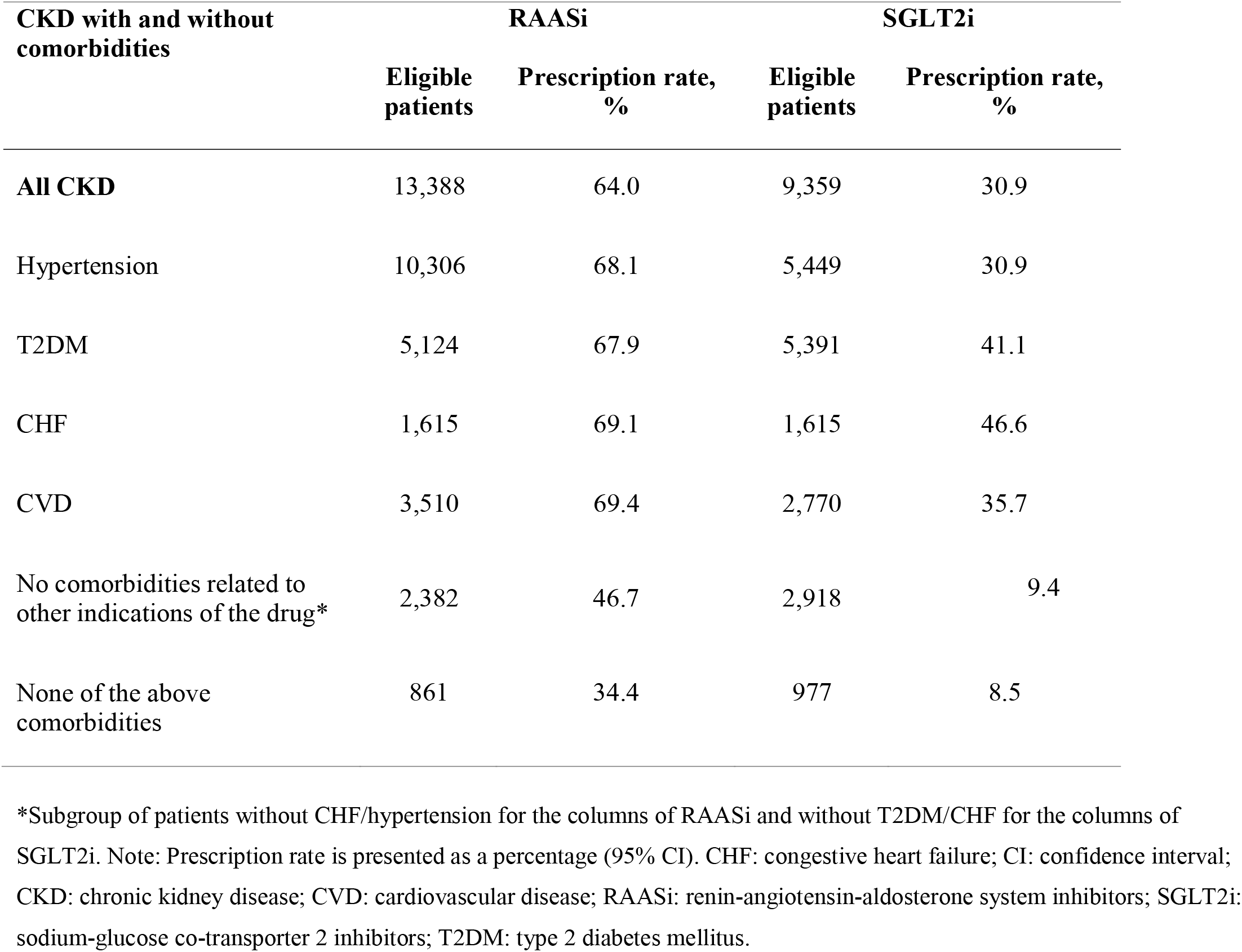
RAASi and SGLT2i prescribing rates post CKD indication and confirmed eligibility in laboratory-confirmed CKD patients with and without comorbidities.

| CKD with and without comorbidities | RAASi |  | SGLT2i |  |
| --- | --- | --- | --- | --- |
|  | Eligible patients | Prescription rate, % | Eligible patients | Prescription rate, % |
| All CKD | 13,388 | 64.0 | 9,359 | 30.9 |
| Hypertension | 10,306 | 68.1 | 5,449 | 30.9 |
| T2DM | 5,124 | 67.9 | 5,391 | 41.1 |
| CHF | 1,615 | 69.1 | 1,615 | 46.6 |
| CVD | 3,510 | 69.4 | 2,770 | 35.7 |
| No comorbidities related to other indications of the drug* | 2,382 | 46.7 | 2,918 | 9.4 |
| None of the above comorbidities | 861 | 34.4 | 977 | 8.5 |

\*Subgroup of patients without CHF/hypertension for the columns of RAASi and without T2DM/CHF for the columns of SGLT2i. Note: Prescription rate is presented as a percentage (95% CI). CHF: congestive heart failure; CI: confidence interval; CKD: chronic kidney disease; CVD: cardiovascular disease; RAASi: renin-angiotensin-aldosterone system inhibitors; SGLT2i: sodium-glucose co-transporter 2 inhibitors; T2DM: type 2 diabetes mellitus.

Among RAASi-eligible patients, 64.8% had evidence of RAASi prescribing prior to their eligibility date. After becoming eligible, an additional 10.0% had a prescription recorded, including 4.4% within 6 months, 1.4% between 6 and 12 months, 2.0% between 12 and 24 months, and 2.2% more than 24 months after eligibility. Nonetheless, 25.2% had no recorded prescription. For SGLT2i, 15.8% had received treatment before eligibility, and an additional 17.6% initiated treatment afterwards, including 4.8% within 6 months, 2.8% between 6 and 12 months, 4.2% between 12 and 24 months, and 5.9% more than 24 months after eligibility. Despite this additional uptake, 66.6% of SGLT2i-eligible patients had no recorded prescription following eligibility, indicating substantial underutilisation (Supplementary Table 5).

## Discussion

### Summary

This study presents a population-level analysis of prevalence, monitoring, and use of GDMT among 19,382 adults with CKD identified in Irish primary care. The cohort had a median age of 76 years, was predominantly female and most were in CKD stage 3 (60.7%). The most common comorbidities were hypertension (53.2%) and T2DM (30.9%).

The age- and sex-standardised CKD prevalence estimate was 5.4% in adults ≥20 years. Among adults ≥50 years, the prevalence under the Expanded definition was 13.0% in CRADLE primary care EHR setting, compared with 15.6% in TILDA.^10^ Although methodologies differed, estimates were closely aligned at ages 50–79 years, with the main divergence among those ≥80 years.

Gaps were identified in monitoring practices. Overall, 66% of CKD-laboratory-confirmed patients underwent follow-up eGFR testing within 12 months of CKD identification. In contrast, uACR monitoring was much less frequent, occurring in <15% of patients within 12 months, and decreasing to 6.6% among those without T2DM. Follow-up eGFR testing rates were similar between patients with and without T2DM, whereas uACR testing was far lower in those without T2DM (34.2% vs. 6.6% in those without T2DM). The low rate of SGLT2i prescribing in non-diabetic patients with CKD supports the need for a dedicated national management program for CKD in Ireland. Without a clear focus on CKD in primary care, the uptake of guideline-directed therapies that slow disease progression and reduce cardiovascular risk is unlikely to be optimised.

Substantial gaps were observed in the use of GDMT, particularly SGLT2i. Among eligible patients, 64.0% received RAASi prescription on or after becoming eligible, compared with only 30.9% for SGLT2i, while no prescriptions for nsMRAs were identified. Prescribing rates for RAASi and SGLT2i were consistently higher among patients with comorbid conditions (T2DM, CHF, and CVD), suggesting that treatment uptake may be driven in part by indications beyond CKD. Although many patients had initiated RAASi before meeting eligibility, one in four had no recorded RAASi prescription after becoming eligible. Notably, two-thirds of patients eligible for SGLT2i had no prescription after meeting treatment criteria, highlighting a considerable implementation gap between evidence-based recommendations and routine clinical practice. These findings suggest that despite the established cardiorenal benefits of contemporary CKD therapies, substantial opportunities remain to improve their identification, initiation, and optimisation in eligible patients, particularly for agents such as SGLT2i.

### Strengths and limitations

This is the largest assessment of CKD burden and management conducted in Irish primary care and the first to use EHR data. While not a direct measure of population prevalence, it provides a practical and policy□relevant view of CKD in primary healthcare. The findings show that CKD is common but often undercoded, highlighting opportunities to improve identification, monitoring, and use of GDMT.^10^

The study benefits from extensive longitudinal data, allowing assessment of coding, monitoring, and prescribing over time. However, there are several limitations. CKD identification relied mainly on eGFR, with limited uACR testing, which may underestimate early-stage disease. Coding inconsistencies mean that some clinically recognised CKD may remain uncoded. Similarly, comorbidities and prescribing patterns were based on recorded data and may not fully reflect true clinical practice. Potential bias may arise from reliance on routinely collected data and laboratory testing, which may miss untested individuals and underestimate prevalence. To address this, sensitivity analyses were performed using alternative CKD definitions and tested-population denominators. Comorbidities were identified from coded diagnoses and may underestimate the true burden of disease. Prescribing should be interpreted in the context of evolving guidelines. Non-prescribing may also reflect contraindications, intolerance, patient preference, or other clinical considerations not captured in routine data. Finally, the findings on CKD monitoring and treatment reflect implementation within the primary care setting and may not be generalisable to secondary care.

Overall, the findings highlight opportunities to strengthen CKD detection, and management in primary care, while also underscoring the challenges of using routine data to capture the full burden of disease.

### Comparison with existing literature

CKD prevalence estimates vary widely across Europe, from 3.3% (Norway) to 17.3% (northeast Germany) for stages 1–5, with that for stages 3–5 from 1.0% (central Italy) to 5.9% (northeast Germany), reflecting disparities in detection and monitoring.^22^ Underdiagnosis remains a concern: German data indicate that only 47% of patients meeting CKD criteria based on laboratory measurements had a documented diagnosis.^17^

Our findings of substantial gaps in CKD monitoring and coding in Irish primary care are consistent with patterns reported across Europe. Consistent with our finding that limited uACR testing may underestimate CKD prevalence, a population-based cohort study in France found that most cases were identified through increased albuminuria rather than reduced eGFR, concluding that routine albuminuria assessment is essential for early detection and preventive treatment, particularly in high-risk populations with comorbidities (hypertension, T2DM, CVD).^21^

CKD prevalence estimates from CRADLE compare favourably with findings from previously published studies from Ireland. The age- and sex-standardised CKD prevalence was 5.36% in adults ≥20 years, compared with 4.5% in Stack et al..^12^ Similarly, the CKD prevalence in individuals ≥ 50 was comparable to estimates in TILDA when similar definition was used (13.0% vs 15.6% respectively).^10^ The gap with TILDA emerged mainly in ≥80 years, consistent with greater detection of asymptomatic laboratory-defined CKD in protocol-driven testing (Supplementary Figure 2). The CKD stage distribution is concordant with previous Irish studies.^10,12^

These findings highlight important opportunities to strengthen the consistency in both CKD coding and monitoring— particularly uACR testing—in primary care, in order to support timely risk stratification and appropriate initiation of GDMTs. Targeted CKD screening in high-risk adults—those ≥45 years with comorbidities as T2DM, hypertension, heart failure, or a history of myocardial infarction or stroke—combined with improved adherence to GDMTs, could substantially reduce dialysis initiation, cardiovascular events, mortality, and kidney replacement costs. These benefits come with a modest increase in early-stage CKD prevalence, reflecting earlier diagnosis and longer survival.^19^

### Implications for research and practice

These findings highlight the substantial burden of CKD and important gaps in CKD management within Irish primary care, particularly in the diagnosis, monitoring, and use of GDMTs. Limited testing and follow-up monitoring of uACR, together with low uptake of RAASi and particularly SGLT2i are missed opportunities for timely risk stratification and initiation of evidence-based treatment. Monitoring and prescribing practices appear to be driven more by comorbidities than by CKD itself, potentially delaying optimal kidney and cardiovascular risk management Collectively, these findings underscore the need for the rollout of a national framework with embedded CKD pathways to support timely identification, monitoring, and management of patients across the CKD care continuum.

## Data Availability

The datasets analysed during the current study are not publicly available. Access to the data may be available from the corresponding author on reasonable request and with permission from the relevant data custodians, subject to applicable governance and approval processes.

## Acknowledgments

The authors thank Sarah Cosgrave and Kushagra Rajput (Centric Health IT) and Sam McGlynn and Conor Murphy (ARQ Research Network) for their support of this work. Finally, we thank the staff and patients of Centric Health, whose participation makes this research possible. All the authors meet criteria for authorship as recommended by the ICMJE. The authors did not receive payment related to the development of the manuscript. Irene Fernández Meco (BPharm) and David Herrero (PhD) provided medical writing support which was contracted by and funded by Boehringer Ingelheim. Boehringer Ingelheim was given the opportunity to review the manuscript for medical and scientific accuracy as well as intellectual property considerations. The study was funded and supported by Boehringer Ingelheim. The funder contributed to the study design but did not have access to the data and was not involved in the data analyses.

## Ethical approval

Ethical approval was granted by the Irish College of General Practitioners Research Ethics Committee (Reference: 2682). The study was conducted under the Health Research Regulations 2018 (S.I. No. 314/2018) provisions for retrospective chart review, requiring analysis within the healthcare organisation’s secure environment and precluding external sharing of individual-level data.

## Abbreviations

CHF: congestive heart failure
CI: confidence interval
CKD: chronic kidney disease
CRADLE: clinical research and data-driven learning environment
CVD: cardiovascular disease
eGFR: estimated glomerular filtration rate
EHR: electronic health records
GDMT: guideline-directed medical therapy
ICDD10: International Statistical Classification of Diseases and Related Health Problems, Tenth Revision
ICPCD2: International Classification of Primary Care, Second Edition
IQR: interquartile range
KDIGO: Kidney Disease Improving Global Outcomes
RAASi: renin-angiotensin-aldosterone inhibitors
SGLT2i: sodium-glucose cotransporter 2 inhibitors
TILDA: The Irish Longitudinal Study on Ageing
T2DM: type 2 diabetes mellitus
uACR: urine albumin-to-creatinine ratio

