## Supplemental file for "Prevalence and Management of Chronic Kidney Disease in Irish Primary Care Network"

**Corresponding author:** Benjamin M. Jacob

ORCID: <https://orcid.org/0000-0003-1119-064X>

**Supplementary Tables**

- *Supplementary Table 1. Operational definitions of CKD.*

| Definition | Purpose | eGFR criterion | uACR criterion | Another criterion |
| --- | --- | --- | --- | --- |
| Strict | Approximate KDIGO-style chronicity-based ascertainment | Two eGFR values <60 recorded ≥90 days apart within 12 months | One uACR >70, or two uACR values 3–70 recorded ≥90 days apart | None |
| Practical | Primary analysis; reduce under-ascertainment in routine care | As strict definition | As strict definition, or a single uACR >3 | None |
| Expanded | Sensitivity analysis and comparison with TILDA estimates | As practical definition, or a single most recent eGFR <60 without a prior normal eGFR in the preceding 12 months | As practical definition | Coded CKD diagnosis |

eGFR is expressed in mL/min/1.73m² and uACR in mg/mmol. CKD: chronic kidney disease; eGFR: estimated glomerular filtration rate; uACR: urine albumin-to-creatinine ratio.

- *Supplementary Table 2. Code list for coded diseases/comorbidities*

| Disease | ICD codes | ICPC codes |
| --- | --- | --- |
| CKD | N18, N25, I12, E10.2, E11.2 | U88, U90, U98 |
| Hypertension | I10, I11, I15, I12, I13 | K85, K86, K87 |
| T2DM | E11 | T90 |
| CHF | I50, I42.0, I42.9, I11.0, I25.5, I13.0, I13.2 | K77 |
| CVD | I21, I22, I23, I24, I25, I70, I73, I77, I78, I60, I61, I62, I63, I69.0, I69.1, I69.2, I69.3 | K74, K90, K91, K92, K99 |

CHF: congestive heart failure; CKD: chronic kidney disease; CVD: cardiovascular disease; ICD: International Classification of Diseases; ICPC: International Classification of Primary Care; T2DM: type 2 diabetes mellitus.

- *Supplementary Table 3*. *Therapy-specific eligibility definitions.*

| Therapy | Eligibility criteria |
| --- | --- |
| SGLT2i | Laboratory-confirmed CKD with eGFR ≥20 ml/min per 1.73 m2 and either of:   - Type-2 Diabetes - uACR ≥20 mg/mmol or with chronic heart failure regardless of uACR - eGFR <45 ml/min per 1.73 m2 and uACR <20 mg/mmol |
| RAASi | Laboratory-confirmed CKD with   - uACR ≥3 mg/mmol (A2), or - Hypertension/chronic heart failure regardless of uACR |
| nsMRA | Laboratory-confirmed CKD with   - Type-2 Diabetes - eGFR >25 ml/min per 1.73 m2 - uACR >3 mg/mmol |

### *Supplementary Table 4. Age and sex adjusted prevalence by CKD definition*

| Age Group | Gender | Total | Practical CKD | Prevalence (95% CI) | Population | Population Weight | Weighted Prevalence (%) |
| --- | --- | --- | --- | --- | --- | --- | --- |
| 20-29 | Male | 36059 | 39 | 0.11% (0.11–0.11) | 326700.0 | 0.081449 | 0.008809 |
| 20-29 | Female | 42350 | 70 | 0.17% (0.17–0.17) | 315700.0 | 0.078707 | 0.013009 |
| 30-39 | Male | 45616 | 99 | 0.22% (0.22–0.22) | 348500.0 | 0.086884 | 0.018856 |
| 30-39 | Female | 58833 | 130 | 0.22% (0.22–0.22) | 375700.0 | 0.093665 | 0.020697 |
| 40-49 | Male | 48251 | 337 | 0.70% (0.70–0.70) | 402300.0 | 0.100297 | 0.070050 |
| 40-49 | Female | 56123 | 387 | 0.69% (0.69–0.69) | 426300.0 | 0.106280 | 0.073286 |
| 50-59 | Male | 37432 | 843 | 2.25% (2.25–2.25) | 340300.0 | 0.084840 | 0.191066 |
| 50-59 | Female | 40586 | 869 | 2.14% (2.14–2.14) | 347200.0 | 0.086560 | 0.185336 |
| 60-69 | Male | 26148 | 1536 | 5.87% (5.87–5.87) | 267800.0 | 0.066765 | 0.392193 |
| 60-69 | Female | 29296 | 1784 | 6.09% (6.09–6.09) | 278800.0 | 0.069507 | 0.423268 |
| 70-79 | Male | 14073 | 2963 | 21.05% (21.04–21.06) | 185700.0 | 0.046297 | 0.974750 |
| 70-79 | Female | 16196 | 3525 | 21.76% (21.75–21.77) | 198000.0 | 0.049363 | 1.074368 |
| 80+ | Male | 7167 | 2763 | 38.55% (38.54–38.56) | 84200.0 | 0.020992 | 0.809267 |
| 80+ | Female | 10404 | 4037 | 38.80% (38.79–38.81) | 113900.0 | 0.028396 | 1.101840 |

CI: confidence interval; CKD: chronic kidney disease

Age and sex adjusted prevalence with 95% confidence intervals and standardised weighted prevalence.

- *Supplementary Table 5. Timing of evidence-based therapy initiation following confirmed post-CKD eligibility.*

| Drug | Pre-CKD Eligibility (%) | <6 months (%) | 6-12  months (%) | 12-24  months (%) | >24  months (%) | Never (%) |
| --- | --- | --- | --- | --- | --- | --- |
| Eligible for RAASi | 64.77 | 4.38 | 1.43 | 1.98 | 2.23 | 25.19 |
| Eligible for SGLT2i | 15.75 | 4.77 | 2.76 | 4.16 | 5.93 | 66.64 |

CKD: chronic kidney disease; RAASi: renin-angiotensin-aldosterone inhibitors; SGLT2i: sodium-glucose cotransporter 2 inhibitors

**Supplementary Figures**

- *Supplementary Figure 1. Inclusion flowchart for the overall study population*.

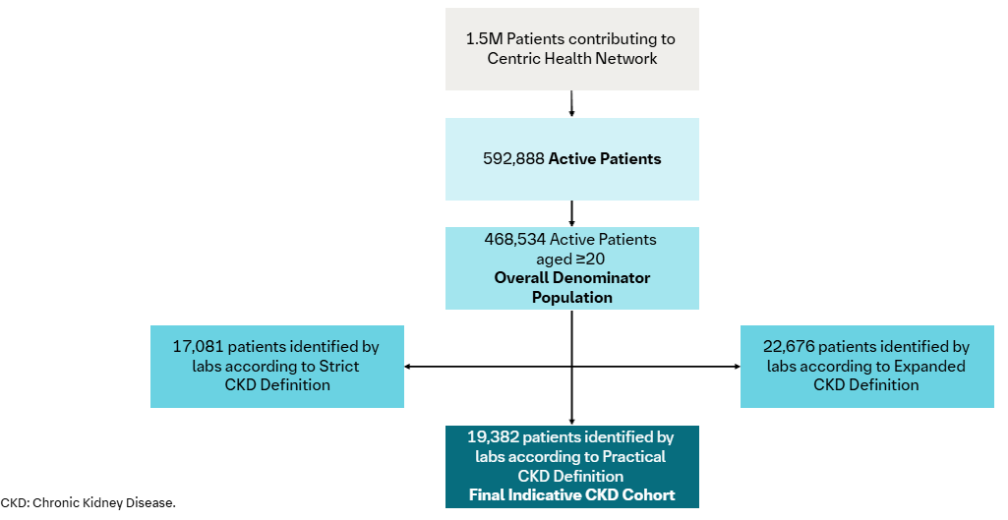

- *Supplementary Figure 2. Age-standardised CKD prevalence stratified by sex: comparison between this study and the TILDA cohort.*

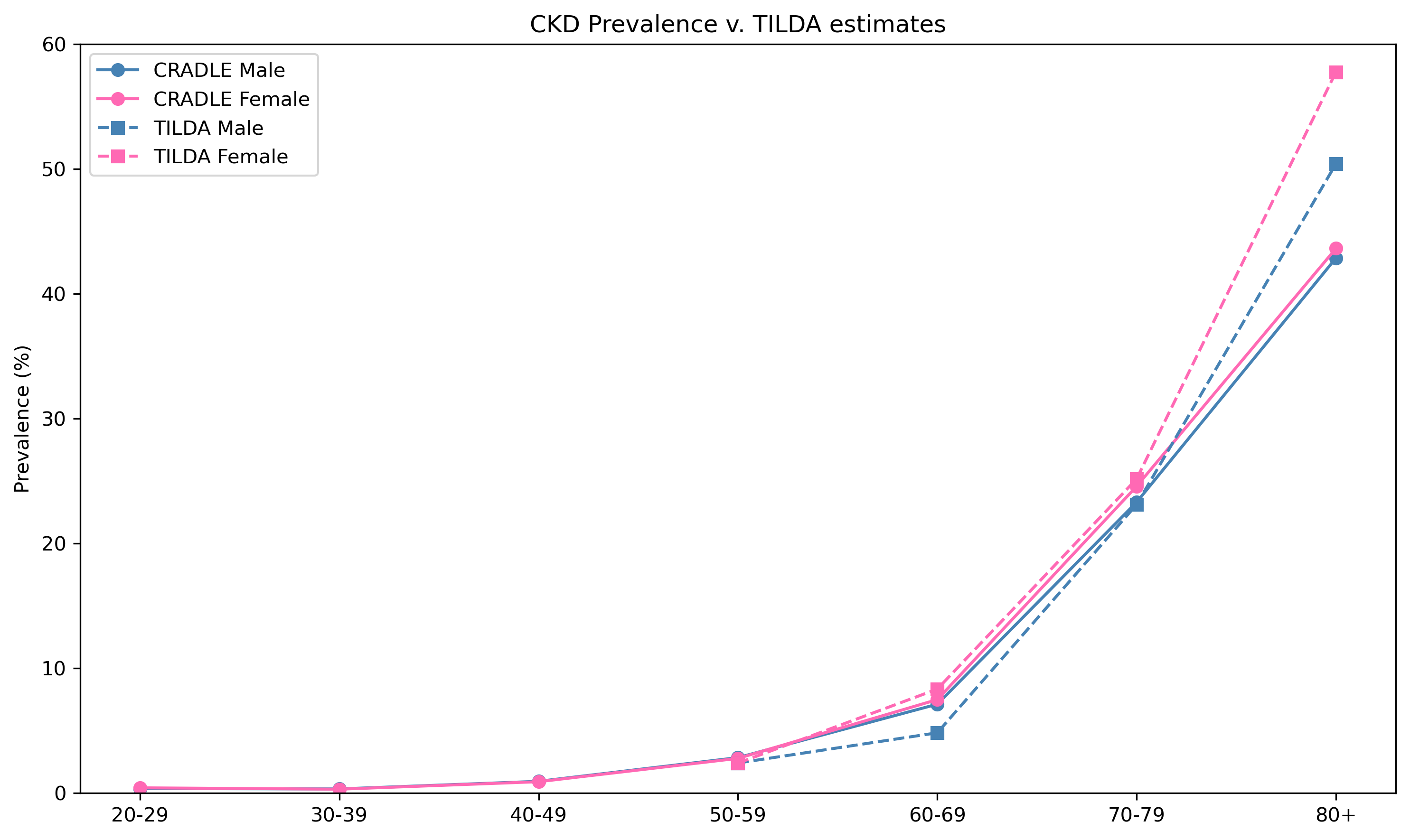

CKD: chronic kidney disease; CRADLE: Clinical Research and Data-Driven Learning Environment; TILDA: The Irish Longitudinal Study on Ageing.

CKD prevalence compared by age and sex subgroups with the estimates in TILDA study, under the same expanded definition.
